# Detection of rare mutations, copy number variation, and DNA methylation in the same template DNA molecules

**DOI:** 10.1101/2022.12.06.22283116

**Authors:** Yuxuan Wang, Christopher Douville, Joshua D. Cohen, Austin Mattox, Sam Curtis, Natalie Silliman, Maria Popoli, Janine Ptak, Lisa Dobbyn, Nadine Nehme, Jonathan C. Dudley, Mahmoud Summers, Ming Zhang, Chetan Bettegowda, Nickolas Papadopoulos, Kenneth W. Kinzler, Bert Vogelstein

## Abstract

The analysis of cell-free DNA (cfDNA) from plasma offers great promise for the earlier detection of cancer. At present, changes in DNA sequence, methylation, or copy number are the most sensitive ways to detect the presence of cancer. To further increase the sensitivity of such assays with limited amounts of sample, it would be useful to be able to evaluate the same template molecules for all these changes. Here we report an approach, called MethylSaferSeqS, that achieves this goal, and can be applied to any standard library preparation method suitable for massively parallel sequencing. The innovative step was to copy both strands of each DNA-barcoded molecule with a primer that allows the subsequent separation of the original strands (retaining their 5-methylcytosine residues) from the copied strands (in which the 5-methylcytosine residues are replaced with unmodified cytosine residues). The epigenetic and genetic alterations present in the DNA molecules can then be obtained from the original and copied strands, respectively. We applied this approach to plasma from 265 individuals, including 198 with cancers of the pancreas, ovary, lung and colon, and found the expected patterns of mutations, copy number alterations, and methylation. Furthermore, we could determine which original template DNA molecules were methylated and/or mutated. MethylSaferSeqS should be useful for addressing a variety of questions relating genetics and epigenetics in the future.

## INTRODUCTION

Detecting cancers at a stage when they can still be cured is one of the major objectives of current cancer research. Assays based on the detection of genetic or epigenetic alterations of DNA, chromosome copy number changes, or changes in the abundance of specific proteins or RNA molecules are being explored for this purpose. Among them, changes in DNA are the most sensitive and specific, with the capacity to identify one altered DNA allele of a given gene mixed with more than 100,000 normal alleles of the same gene(1). To detect such rare alleles requires the purification of DNA from multiple milliliters (mL) of plasma, as 1 mL of plasma contains an average of only ∼1000 to 2000 alleles of each gene. Past studies have shown that increases in sensitivity can be obtained by combining assays for DNA alterations and changes in protein abundance(2, 3). It would be ideal if epigenetic changes could also be assessed in the same cfDNA molecules used to detect genetic changes, thereby reducing the total volume of plasma required from the patient.

The most commonly used method for investigating the epigenetic status of DNA employs bisulfite-mediated deamination to convert unmethylated deoxycytidine residues to deoxyuridine (4-8). Unfortunately, bisulfite treatment makes it impossible to detect the type of mutation that is most common in cancers, i.e., C to T transitions (9). We here report a technique called MethylSaferSeqS that employs bisulfite-mediated deamination but allows detection of any type of genetic or epigenetic change in DNA, including C to T transitions, in the same DNA template.

## RESULTS

### Overview of MethylSaferSeqS

The most sensitive way to detect rare mutations is through duplex sequencing, i.e., the independent determination of the sequence of both strands of DNA. Errors during DNA purification, library preparation, or sequencing are much more likely to alter a single base, rather than both bases at that position in complementary (Watson and Crick) DNA strands. Thus duplex sequencing can exponentially reduce error rates. Several methods for duplex sequencing have been described(1, 10-15). MethylSaferSeqS can be applied to any of these library preparation methods using the following approach:

1. Copy both strands (Watson and Crick) of each template DNA molecules after adapter ligation.
2. Separate the original DNA strands from the copied strands.
3. Prepare two libraries, one from the original DNA strands (a) and one from the copied (b) strands:
  a. Deaminate unmodified cytosine residues in the original strands using bisulfite, PCR-amplify, and assess epigenetic changes;
  b. PCR-amplify the copied strands and assess genetic changes.

Though these steps are simple in principle and involve only minimal changes to conventional library preparation methods, performing them in a way that preserved the majority of the initial DNA template molecules for both genetic and epigenetic analysis is challenging. The final, optimized protocol is summarized in **Fig. 1**, with further details described below and in Materials and Methods.

**Fig. 1:**
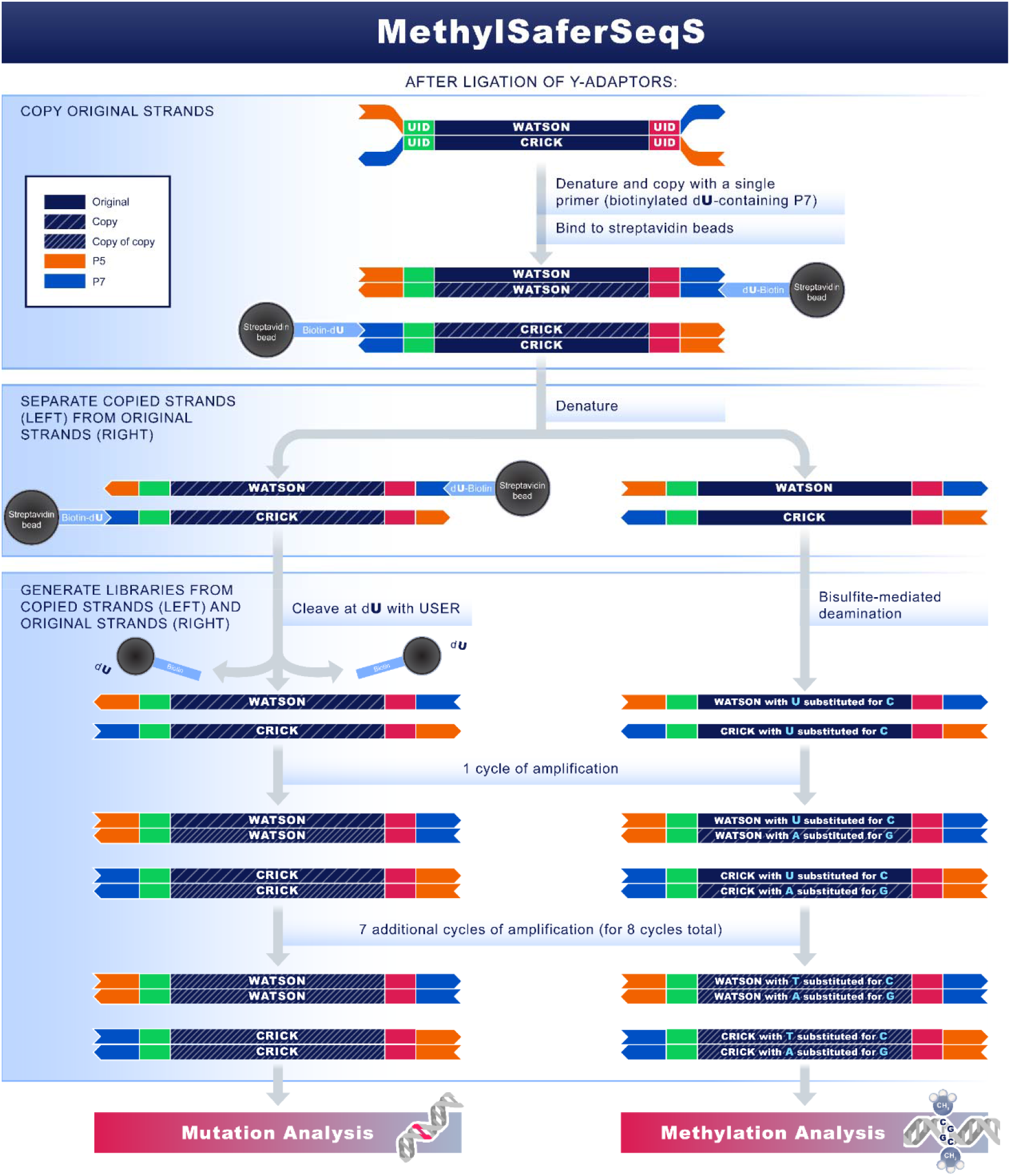
Overview of MethylSaferSeqS. Following ligation of Y-adaptors to each double-stranded DNA molecule during standard library construction, each original strand is copied with a primer that targets the P7 sequence. The primer contains a dual biotin modification and a deoxyuridine (dU) base at the 5’ end. The 5’ biotins allow the copied strand (still hybridized to the original strand) to bind to streptavidin beads. After binding, the original strands are separated from the beads via heat denaturation. The remaining, copied strand is then removed from the beads via cleavage of the deoxyuridine residue at the 5’ end of the primer. This process yields two libraries, one from the copied strands (left) used for mutation analysis, the other from original strands (right) used for methylation analysis. UID: Unique identifier. USER: Uracil DNA glycosylase and the DNA glycosylase-lyase Endonuclease VIII.

### Principles

The first principle involves modifications to adaptor sequences to ensure compatibility with standard library preparation and sequencing methods. The adapters appended to the template molecules must be modified such that bisulfite-mediated deamination does not change the adapter sequences in a way that would make them incompatible with other components of standard, commercially available library generation and sequencing processes. Our preferred change was to substitute cytosine (C) residues with 5-methylcytosines (5-meC’s) in the adapter sequences. Adapters could also be changed in other ways for the same purpose, such as using only A, T, and G’s without any C’s.

The second principle is embodied in the statement “every molecule is sacred”(16). Because abnormal genetic or epigenetic changes are expected to be very rare in certain cfDNA samples, avoiding losses of these alleles during library preparation is critical. The SaferSeqS library preparation method yields a higher fraction of template molecules than most other library preparation methods(1), and was therefore used as the base for the MethylSaferSeqS protocol. SaferSeqS also facilitates duplex sequencing, which is key to the reliable detection of rare mutations. Though the base workflow described here for MethylSaferSeqS is that used for SaferSeqS, the protocol can be easily adapted to any library preparation method.

### Step 1: Copy template molecules

The addition of this step represents the major deviation from conventional library preparations. In conventional library preparations, once adapters are ligated to the DNA templates, the library is amplified using PCR to obtain sufficient DNA for target enrichment or for other purposes. Once amplification is performed, however, all covalent modifications of the DNA templates, such as methylation or hydroxymethylation, are lost and replaced by unmodified deoxyribonucleic acid residues during the copying process. In MethylSaferSeqS, the two DNA strands are heat denatured as in a typical PCR step, but then each strand is copied with a single primer containing a dual biotin modification and a deoxyuridine (dU) at its 5’ end (described in Step 2 below). Because only one rather than two primers per reaction are used for this step, exponential amplification does not occur. Following this copying, there are *n*+1 strands rather than 2^n^ strands (*n* = the number of PCR cycles performed): the original DNA strand (containing covalent modifications such as methylation) and *n* copied strands (devoid of such modifications).

During the initial development of the MethylSaferSeqS protocol, we used only one round of denaturation and copying with the biotinylated, dU-containing primer. In later stages, we used two or three sequential rounds of denaturing and copying, always with a single primer rather than two primers to avoid exponential amplification. The reason for using more than one cycle was to increase yield, given the importance of recovering as many molecules as possible. With three cycles, essentially all of the template molecules that were successfully recovered in the base protocol (SaferSeqS) were also recovered with MethylSaferSeqS, as assessed by duplex sequencing (**Fig. 2**). With one and two cycles, the recoveries were 37% and 81%, respectively.

**Fig. 2:**
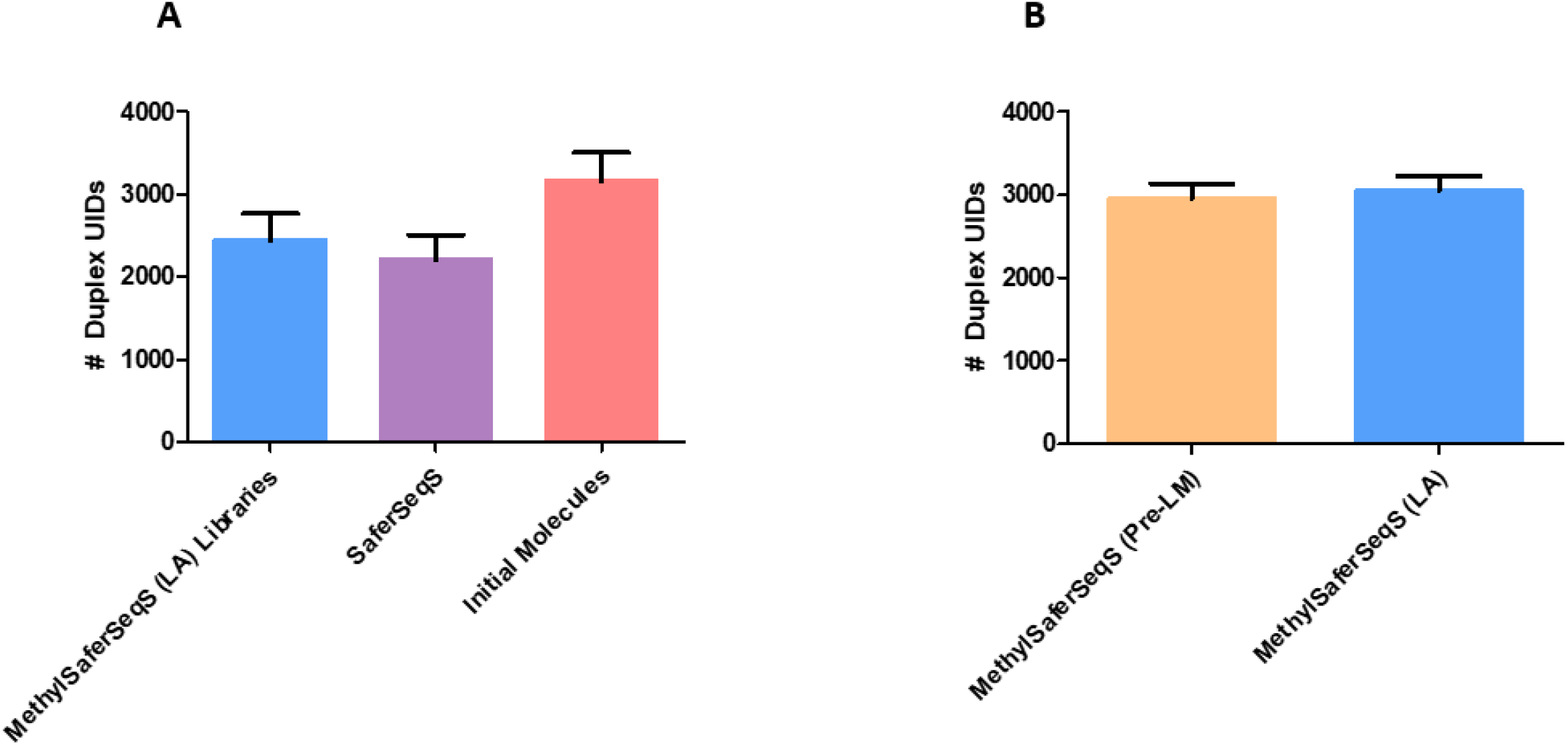
Duplex Recovery of MethylSaferSeqS compared to standard library construction approach (SaferSeqS) and the number of original molecules. The numbers of distinct molecules present in MethylSaferSeqS and SaferSeqS were assessed through duplex sequencing after target enrichment through amplification of 48 genomic regions of interest. Each distinct molecule is tagged with a unique identifier (UID), which allows for precise quantification of the number of initial DNA molecules. A duplex UID is defined as an UID that was found in both the Watson and Crick strands of the molecule. **(A)** The average duplex recoveries were 77% and 70% for MethylSaferSeqS (LA) and SaferSeqS libraries, respectively, compared to the total number of initial DNA molecules. **(B)** Within the MethylSaferSeqS libraries, similar numbers of distinct molecules were partitioned for methylation analysis prior to bisulfite treatment (pre-LM) and mutation detection (LA).

### Step 2: Separating the copied strands from the original strands

The primer used for copying contained a dual biotin modification and a dU at the 5’ end as described above. The 5’ dual biotin modification allows the copied strand (still hybridized to the original strand) to bind to streptavidin beads and ensures that this interaction is not disrupted by heat. After binding, the original strands are separated from the beads and bound copied strands via heat denaturation. Lastly, the remaining, copied strand is removed from the beads by cleavage at the dU residue, using uracil DNA glycosylase and DNA glycosylase-lyase Endonuclease VIII. The removal of DNA templates from beads was done because we found that for subsequent library construction, amplification in solution was more efficient than amplification on beads.

Many variations of Steps 1 and 2 were evaluated during the development of MethylSaferSeqS. These included singly-biotinylated rather than doubly-biotinylated primers for copying the original template, various numbers of deoxyuridines (from zero to six) at various positions in the primers, various conditions for binding to streptavidin beads, temperatures, EDTA concentrations, and pH for separating the original strands and copied strands from the beads. Though all variations recovered the desired molecules to some degree, the detailed protocol described in Methods performed best of those tested.

### Step 3a. Use bisulfite to deaminate the C’s in the original strands

Bisulfite-mediated deamination results in cleavage of DNA (17, 18). Longer reaction times and higher temperatures in the presence of bisulfite results in higher conversion rates of unmethylated cytosine to uracil, but lower DNA recovery. Given that many of the aberrantly methylated molecules in plasma from early stage cancer patients are expected to be present at low frequencies, we attempted to find conditions that would lead to reasonable DNA yields while maintaining an acceptable conversion efficiency. Through trial and error, we found that bisulfite treatment at 50°C for 3 hours yielded results that were optimal for our purposes. This preserved the majority (∼70%) of the input template molecules with 80% of unmethylated C’s converted to U’s. This conversion efficiency was adequate for the regions of interest to us – i.e., those containing relatively abundant CpG sites. Greater (>98%) conversion of the unmethylated C’s could be achieved by exposure to bisulfite at 50°C for 16 hours, but this resulted in loss of half of the original DNA template molecules when assessed by the Methods described in Performance Evaluation 2 below.

The amplification of the bisulfite-treated library was performed as in Step 3b below with one important difference: the primers for PCR were shorter, truncated at the 3’ end after a GC dyad. In the copied molecules, all C’s would be converted to T’s following bisulfite treatment, including those in the primer binding sites. Thus only the original molecules that have methylated adaptors would retain homology to primer sequences and would be selectively amplified during PCR. The reason for the primer change from the original library amplification primers was to minimize the effects of any unintended copied strands that end up in the libraries of the original DNA strands during Step 2. There are two conceivable ways for copied strands to be carried over into the library of original molecules. First, although only one primer was used in Step 2, any contaminating adapter-derived sequences from prior ligation steps could act as primers to produce non-biotinylated copies of the original strand. Because these copies would no longer be methylated at CpG sites, they could artificially decrease the apparent fraction of methylated CpG sites in libraries from original template strands. Second, although the use of dual rather than single biotin should prevent dissociation of biotinylated copied strands from streptavidin during heating(19), any small fraction of copied strands that also come off the streptavidin beads during heat denaturation would contaminate the library with original molecules. Of several primers tested to specifically amplify the original molecules following bisulfite conversion, the one employed in the final protocol (**Table S1**; see Methods) performed best in preserving the expected methylation patterns, as described in Performance Evaluation 5 below.

### Step 3b: PCR-amplify the copied strands

This step was performed identically to that of the basic SaferSeqS protocol(1) after the copied molecules are cleaved from the streptavidin beads described in Step 2. In this step, primers targeting the adaptor sequences at the ends of each DNA fragment are used for amplification (**Table S1**).

At the end of these three steps, the procedure yields two libraries: One from copied molecules for assessment of genetic alterations such as mutations and copy number changes, abbreviated “LA” (Step 3b); and one from original molecules for assessment of DNA methylation, abbreviated “LM” (Step 3a).

### Performance Evaluation # 1: What fraction of the original template molecules are represented in the libraries?

Because each original template molecule has a unique molecular barcode (unique Identifier, abbreviated UID), we could estimate the fraction of template molecules recovered in the LA and LM libraries through amplification of the libraries with specific primers for genomic regions of interest. We used 48 regions that were commonly mutated in cancers as previously described(1) to evaluate libraries generated from the plasma DNA of 7 healthy individuals (**Table S2**). This procedure uses a hemi-nested PCR to enrich the regions of interest. Because evaluation of both strands of the same molecules are required for the most sensitive and accurate detection of mutations, the recovery of duplex UIDs was used as a metric for the efficiency of library construction. Each of the cfDNA samples was divided in two equal aliquots: one aliquot was used to construct MethylSaferSeqS libraries and the other to construct standard SaferSeqS libraries. As shown in **Fig. 2A**, the number of duplex UIDs recovered in the LA form of MethylSaferSeqS libraries were comparable to that in the standard SaferSeqS libraries. The average recovery of duplex UIDs for the MethylSaferSeqS LA libraries was 78% (interquartile range (IQR): 68 to 89%) of the initial DNA template molecules in the cfDNA. This recovery represents a lower bound because it does not take into account losses of template molecules during the hemi-nested PCR enrichment step used for mutation analysis.

To estimate the fraction of templates recovered during preparation of the LM libraries, we generated “Pre-LM Libraries” from the DNA strands eluted from Streptavidin beads after heat denaturation but before bisulfite conversion in Step 2. In parallel we amplified the DNA from the copied strands which remained bound to the beads (i.e., the LA libraries) using the same primers. The duplex UIDs recovered from the pre-LM libraries averaged 97% (IQR 94% to 100%) of those recovered in the LA libraries (**Fig. 2B**).

We did not evaluate the fraction of templates remaining after bisulfite conversion in the LM libraries using this hemi-nested PCR-based approach. After bisulfite treatment, the Watson and Crick strands are no longer complementary, making primer design targeting the same region of both strands to accurately estimate duplex recovery difficult, and duplexes were not required for the methylation experiments described below.

### Performance Evaluation # 2: Are the same template molecules represented in the LA and LM libraries?

As the majority of the original plasma template molecules were present in both the LA and pre-LM libraries (**Fig. 2**), there must be considerable overlap between the template molecules in these two libraries based on arithmetic. To quantify the degree of overlap, we performed experiments similar to those described above, but evaluated the fraction of molecules that were shared between the two libraries as determined by their common UIDs (**Fig. 3**) rather than estimating the total number of template molecules recovered in the libraries.

**Fig. 3:**
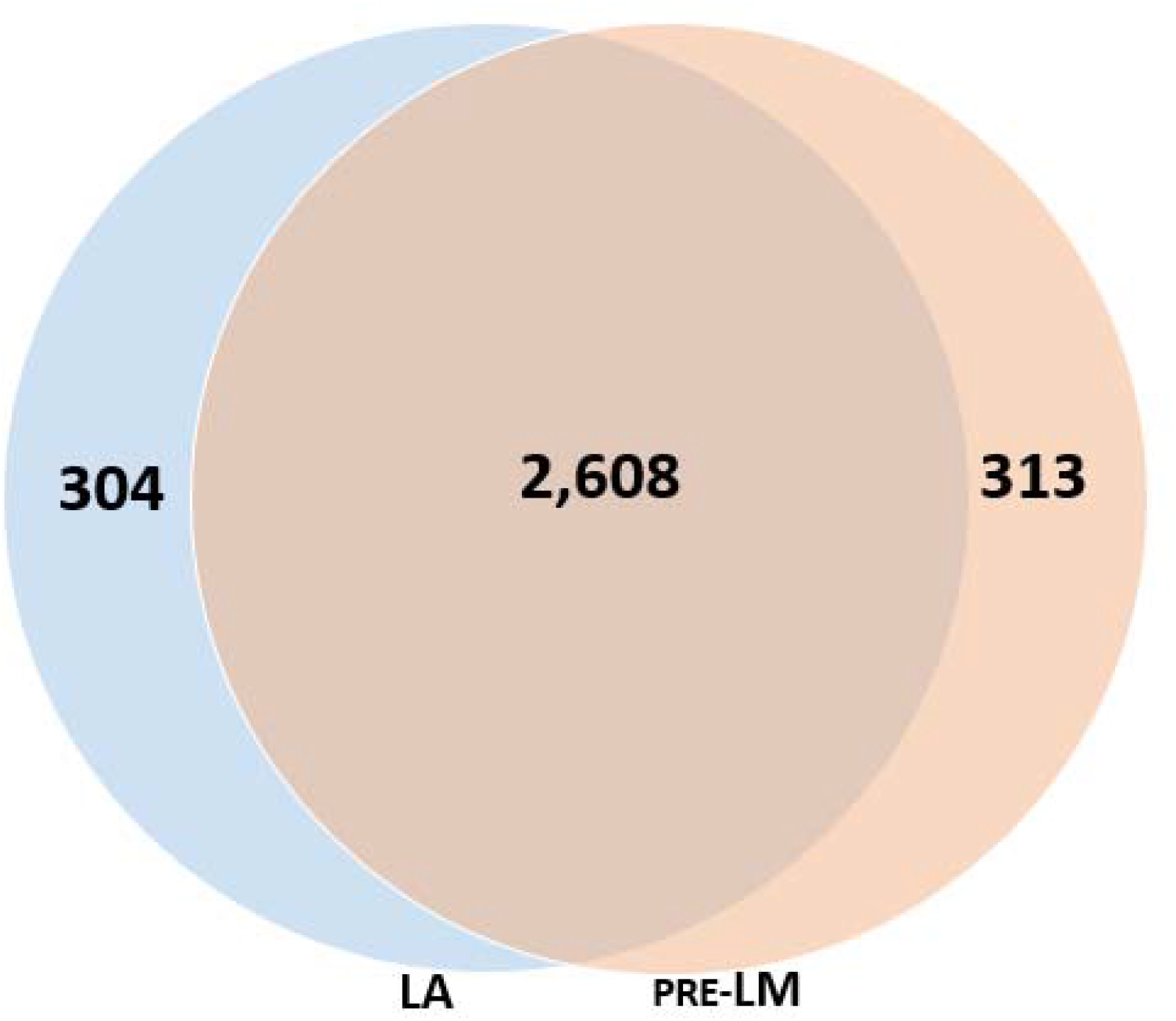
DNA molecules represented in both MethylSaferSeqS LA and pre-LM libraries. Venn diagram depicting the number of DNA molecules represented in both the MethylSaferSeqS LA and pre-LM libraries, as estimated based on the numbers of common duplex UIDs.

Analogous to the experiment depicted in **Fig. 2B**, we first evaluated the shared molecules in the pre-LM libraries made prior to bisulfite-mediated deamination. An average of 2,921 UIDs (IQR: 2,885 to 2,957) were identified in the pre-LM libraries made from the original template strands and 2,912 UIDs (IQR: 2,834 to 2,991) in the LA libraries made from the copied template strands. On average 2,608 UIDs (IQR: 2,553 to 2,662) were present in *both* the pre-LM and LA libraries, which comprise of about 90% of each library. Conversely, an average of 314 UIDs (10.7%, IQR 10.2 to 11.2%) were present *only* in the pre-LM libraries, while 305 (10.5%, IQR: 9.9% to 11%) were present *only* in the LA libraries (**Fig. 3**).

### Performance Evaluation # 3: Can MethylSaferSeqS libraries be used to detect the mutations observed in standard libraries?

Cell-free plasma DNA samples from 19 patients with advanced colorectal cancer (**Table S2**) were divided into two equal aliquots. One aliquot was used to construct MethylSaferSeqS libraries and the other to construct standard SaferSeqS libraries. These libraries were then queried for somatic mutations in 48 genomic regions commonly mutated in these cancers, as described for Performance Test 1. A total of 35 somatic mutations were observed, and of these, 22 (63%) were observed in both the MethylSaferSeqS and SaferSeqS libraries. The MAFs of the mutations found in both libraries were highly concordant (**Fig. 4**; Pearson’s r = 0.9, p<1×10^−9^), with an average MAF of 17.6% (range 0.04% to 79.6%). The remaining 13 (37%) of mutations were found in only one of the two libraries (5 in SaferSeqS only, and 8 in MethylSaferSeqS only) at much lower MAF (average 0.16%; range 0.04% to 0.69%), and all of these were in the stochastic realm as each library generated was from a separate ∼5-mL aliquot of plasma.

**Fig. 4:**
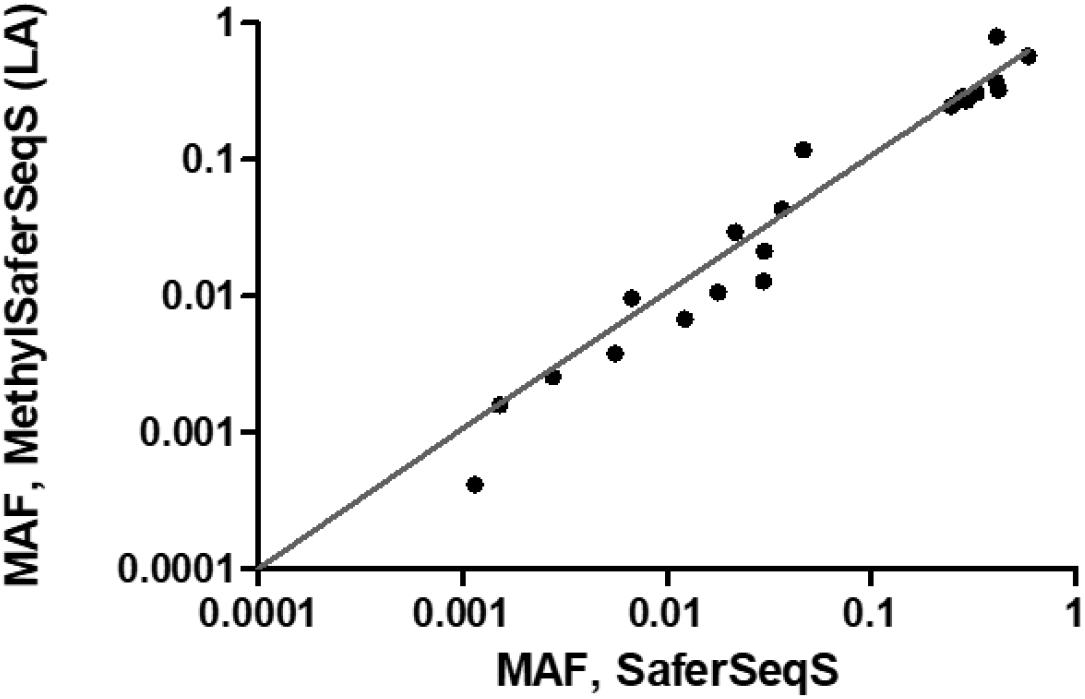
Concordance of mutations and their MAFs identified in cfDNA by MethylSaferSeqS and SaferSeqS. Equal aliquots (∼5ml) of plasma from 19 patients with stage IV colorectal cancer (CRCs) were made into MethylSaferSeqS and SaferSeqS libraries. The MethylSaferSeqS LA and SaferSeqS libraries were assessed for mutations in 48 commonly mutated genomic regions. Twenty-two mutations were identified in both libraries. The mutant allele fraction (MAF) in the MethylSaferSeqS vs. SaferSeqS libraries for these mutations are shown (axes are on logarithmic base 10 scale, range: 0.01% to 100%).

In addition to the 19 patients for whom paired conventional SaferSeqS and MethylSaferSeqS libraries were available, we analyzed MethylSaferSeqS LA libraries from another 246 patients with the same panel of 48 amplicons. In sum, we evaluated MethylSaferSeqS LA libraries from 265 patients, including 198 patients with cancers of the colon, ovaries, pancreas, or lung, as well as 67 healthy individuals, to determine whether the fraction of samples with somatic mutations detectable in the cfDNA conformed to those expected from prior studies. Of the 198 cancer patients, 121 (61%) patients had at least one detectable somatic mutation (**Table 1**). The mutations detected are listed in **Table S3**. As expected, higher fractions of patients with later stage cancers (79% of patients with Stage III or IV cancers) than in earlier stage cancers (49% of patients with Stage I or II cancers, p<1×10^−11^, Binomial test) had detectable mutations. The mutant allele fractions observed were also considerably higher in patients with later stage cancers than early stage cancers (6.6% vs. 1.5%, p<0.005, Student T-test). In the 67 samples from healthy individuals, only one (1.5%) had a detectable mutation in their cfDNA at a low MAF (0.25%; **Table S3**). This particular mutation (*GNAS* p.R201H) is commonly observed in individuals with clonal hematopoiesis of indeterminate potential (CHIP) (20).

**Table 1:** Fraction of patients with detectable mutations in cfDNA using MethylSaferSeqS.

| Cancer Type/Stage | # Patients Total | # Patients<br>Detected (% of<br>Total) | Avg MAF (range), % |
| --- | --- | --- | --- |
| **Lung (Total)** | **32** | **25 (78%)** | **2.21% (0.03% -26.02%)** |
| III | 22 | 20 (91%) | 2.72% (0.03% -26.02%) |
| II | 4 | 2 (50%) | 0.14% (0.13% -0.14%) |
| I | 6 | 3 (50%) | 0.14% (0.09% -0.19%) |
| **Ovarian (Total)** | **37** | **25 (68%)** | **3.61% (0.07% -24.19%)** |
| IV | 2 | 2 (100%) | 1.05% (0.35% -1.74%) |
| III | 17 | 12 (71%) | 2.98% (0.07% -17.12%) |
| II | 5 | 5 (100%) | 7.04% (0.15% -24.19%) |
| I | 5 | 2 (40%) | 1.10% (0.12% -2.08%) |
| NA | 8 | 4 (50%) | 3.77% (0.10% -8.17%) |
| **Pancreas (Total)** | **102** | **49 (48%)** | **0.86% (0.06% -9.12%)** |
| IV | 5 | 5 (100%) | 0.45% (0.26% -0.66%) |
| III | 6 | 1 (17%) | 0.87% (0.87% -0.87%) |
| II | 75 | 37 (49%) | 1.00% (0.07% -9.12%) |
| I | 16 | 6 (38%) | 0.28% (0.06% -0.93%) |
| **CRC (Total)** | **27** | **22 (81%)** | **14.16% (0.07% -58.59%)** |
| IV | 26 | 22 (85%) | 14.16% (0.07% -58.59%) |
| II | 1 | 0 (0%) | 0.00% (0.00% -0.00%) |
| **Healthy Control**<br>**(Total)** | **67** | **1 (1%)** | **0.25% (0.25% -0.25%)** |

### Performance Evaluation # 4: Can MethylSaferSeqS libraries be used to detect copy number alterations observed in standard libraries?

For the analysis of copy number alterations, we used the cfDNA from 17 patients with colorectal cancers (CRC; **Table S2**), from which an aliquot was used to construct MethylSaferSeqS libraries and another to construct standard SaferSeqS libraries, as described above. The libraries were PCR-amplified using primers suitable for loading on an Illumina instrument (**Table S1**), followed by whole genome sequencing to ∼ 0.75x. Nearly identical changes in specific chromosomal regions, as well as breakpoints, were observed in both the SaferSeqS and MethylSaferSeqS LA libraries (**Fig. 5, Table S4**). There were 122 regions with a copy number gain or loss identified in MethylSaferSeqS libraries, of which, 112 (92%) were found with the same change (copy number gain or loss) in the corresponding SaferSeqS libraries. Conversely, there were 132 regions with copy number gain or loss identified in the SaferSeqS libraries, 114 (86%) of these changes were also found in the MethylSaferSeqS libraries. When more stringent criteria were applied to exclude borderline calls (see methods), nearly all remaining calls (98%, or 120 of 122) in the SaferSeqS and MethylSaferSeqS libraries were concordant. No copy number alterations were observed in the SaferSeqS a MethylSaferSeqS libraries derived from cfDNA of the healthy control (**Fig. 5**).

**Fig. 5:**
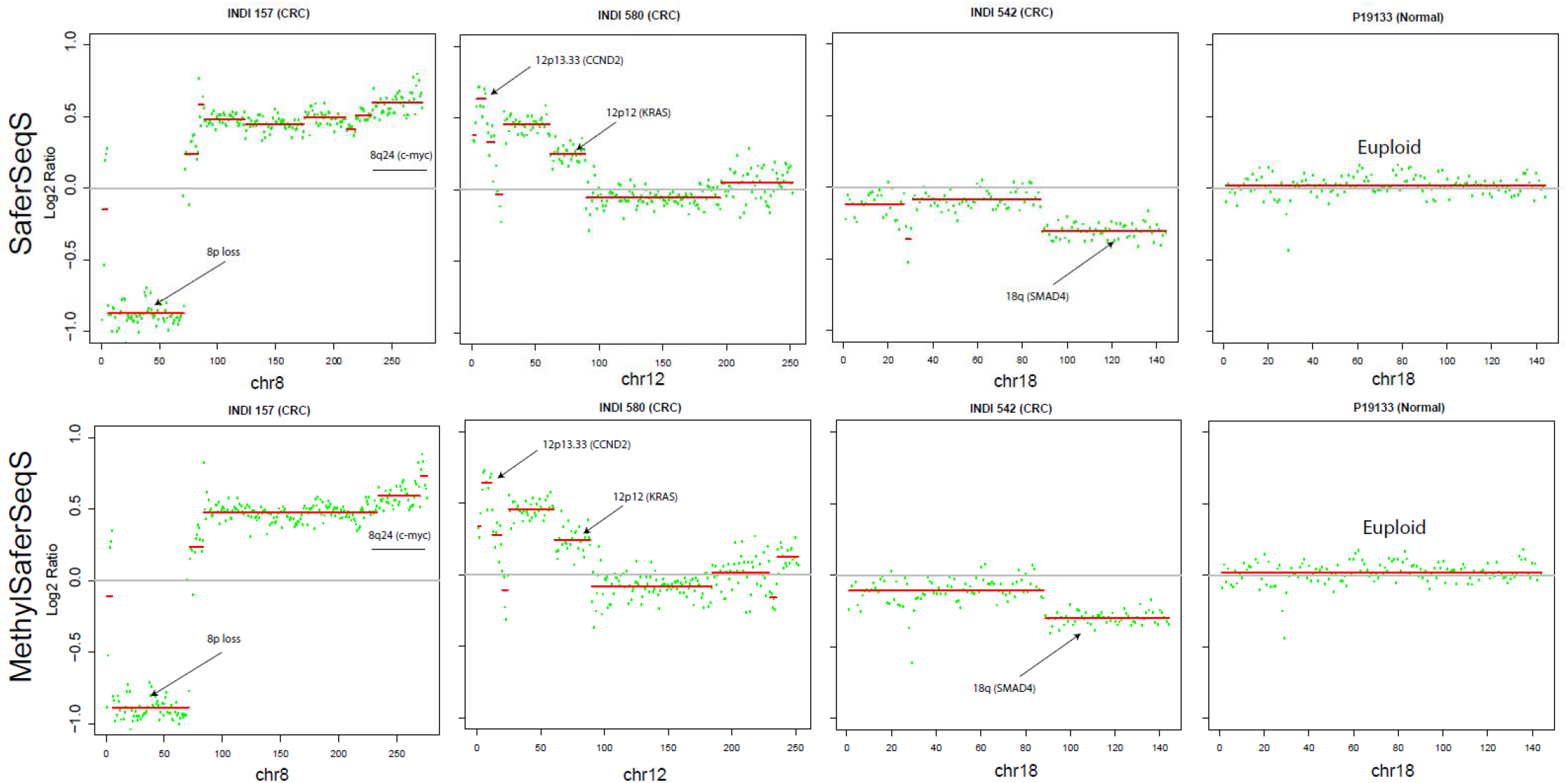
Concordance of copy number variations identified in cfDNA by MethylSaferSeqS and SaferSeqS. MethylSaferSeqS (LA) and SaferSeqS libraries were constructed from equal aliquots of plasma from the patients with advanced colorectal cancer and healthy controls for identification of copy number changes. Representative regions are shown for three patients with colorectal cancer (CRC) and a healthy control. Y-axis represent the number of bins (each bin represents a 500KB genomic region).

### Performance Evaluation # 5: Can MethylSaferSeqS libraries reliably detect DNA methylation?

LM libraries underwent bisulfite-mediated deamination as described in Step 3a above, with average conversion efficiency of 77% (IQR: 75% to 79%). We evaluated the LM libraries in two ways. First, we assessed the contribution of various cell types to cfDNA from healthy individuals. We performed whole genome sequencing to an average depth of ∼ 7 haploid genome equivalents. We then used 5,612 differentially methylated regions (DMR) that were characteristic of given cell types, as previously described (21, 22) This allowed us to define the contributions from the liver, lungs, colon, small intestines, pancreas, adrenal glands, esophagus, heart, brain, T cells, B cells, and neutrophils using quadratic programming in 31 healthy individuals (**Table S5**). In the MethylSaferSeqS libraries, the fraction of template molecules derived from each of the twelve cell types analyzed was similar to expected (**Fig. S1**). For example, leukocytes were by far the most predominant contributors, with average contributions of 68.6% (IQR: 67.2-73.3%), consistent with prior findings(21, 23). Among leukocyte subsets, neutrophils were the greatest contributor, with average contributions of 43.8% (IQR: 40.5%-48.9%). The average neutrophil-to-lymphocyte ratio (NLR) was 1.8 (IQR: 1.6 to 2.1), consistent with the expected ∼2:1 ratio in the circulation of healthy individuals(24). Normal liver was the highest contributor among non-hematopoietic cells, with an average contribution of 8.2% (IQR: 5 to 7.8%), also consistent swith prior findings(21). There were greater than expected fraction of cfDNA (7.5%, IQR 6.4 to 8.6%) identified as derived from the brain. This higher than expected fraction has been observed previously(23), which has been attributed to an artifact of deconvolution algorithms, and less likely, turnover of peripheral neurons.

Second, LM libraries from the cfDNA of cancer patients were PCR-amplified using the primers described in step 3a that were specific for the original molecules after bisulfite conversion (**Table S1**). We then performed whole genome sequencing to an average depth of ∼ 4.1 haploid genome equivalents. To illustrate that MethylSaferSeqS could detect cancer-specific methylation, the data from 11 genomic regions that have previously been reported to be hypermethylated in cancers are recorded in **Table S6** (25-30). The average size of the regions was 114 bp (range 93 to 132 bp), containing an average of 12.6 CpG sites per region (range 7 to 19 bp). This provided an opportunity to discover methylation differences in cfDNA samples in which sufficient amounts of DNA was derived from neoplastic cells. By rough estimation, each marker could be covered in at most 52 distinct DNA molecules (average of 12.6 CpG’s × average coverage of 4.1 per CpG). Thus we applied the panel to plasma samples with mutation MAF>2%, as determined in Performance Evaluation #3, assuming at least one mutant DNA molecule was captured by the panel.

Fractional methylation was calculated for each marker as the ratio of the total number of unconverted cytosines in CpG sites to the total number of CpG sites (See Methods). The fractional methylation was considerably higher in the cfDNA of the 29 cancer patients who had sufficient neoplastic content (MAF>2%) than in the cfDNA samples from the 67 healthy individuals used as controls (34% vs. 5%, p<1×10^−5^, Student’s t-Test; **Table S7**). Moreover, there was a correlation between the mutation MAF found in LA libraries and the fractional methylation in these samples (Pearson’s r=0.9, p<1×10^−9^; **Fig. S2**). Finally, although the markers in our panel were chosen to be cancer-specific rather than tissue-specific, some markers appeared to be specific for certain cancers. For example, the ones reported to be altered in colorectal cancers were more often methylated in the cfDNA from patients with colorectal cancers than in patients with other types of cancers or in healthy individuals (**Fig. 6**).

**Fig. 6:**
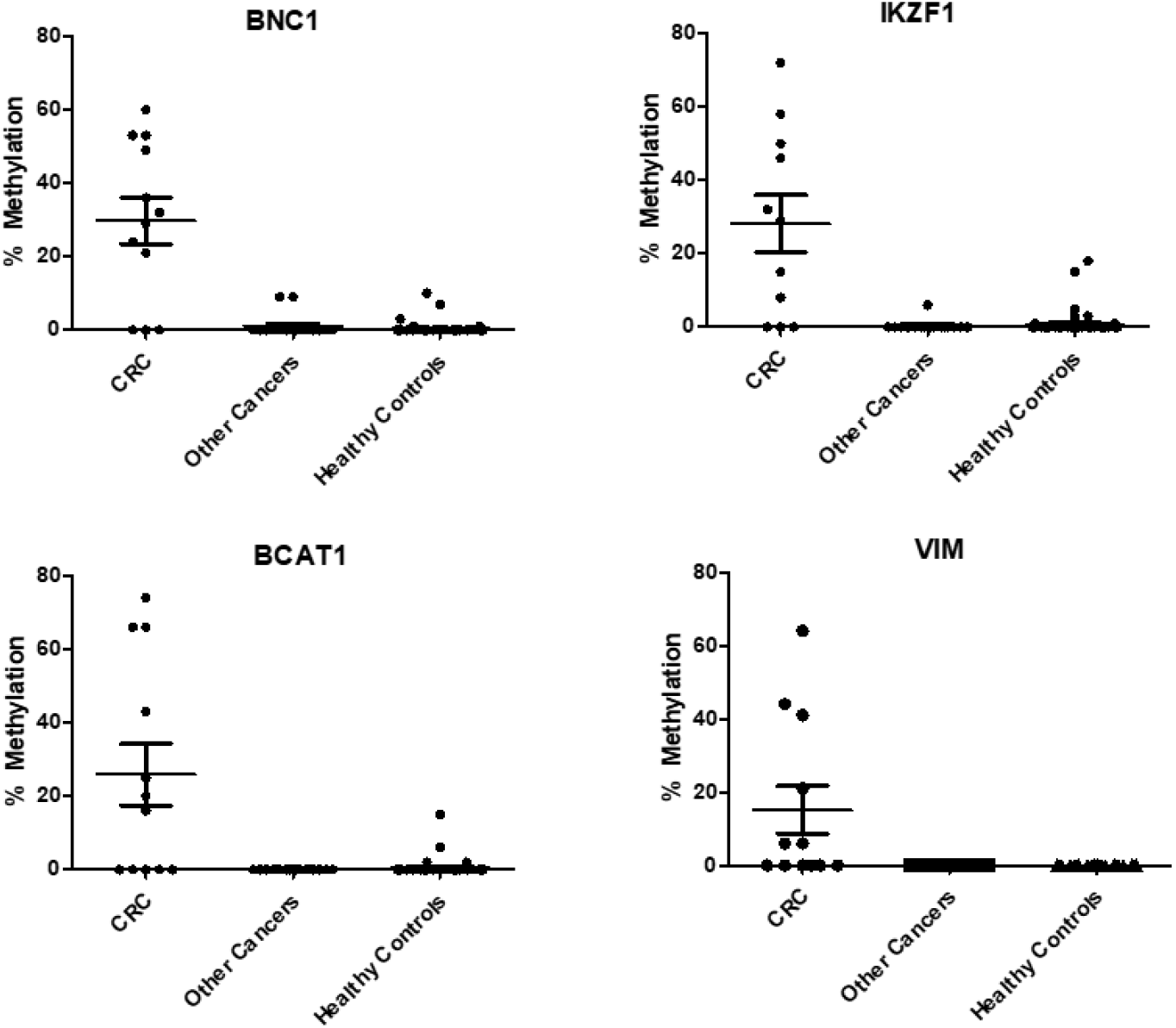
Fraction methylation idenfied in cfDNA samples using markers that are hypermethylated in colorectal cancers (CRCs). MethylSaferSeqS LM libraries from plasma samples of 29 patients with colorectal (CRC), ovarian, pancreatic, and lung cancers that had sufficient neoplastic content (>2% mutant allele frequency based on mutation assay using LA libraries) as well as 67 healthy controls were subjected to whole-genome sequencing. The fractions of methylation in 4 representative markers that are commonly hypermethylated in CRCs are shown.

Finally, we wished to determine whether we could reliably access the methylation status of mutated DNA fragments present in the plasma. For this purpose, we examined two patient samples with mutations that occurred in a CpG dyad that affected the methylation status of the strands (**Fig. 7**). Because the Watson and Crick strands of the original molecules lose complementarity after bisulfite conversion, we used two sets of PCR primers (one for each strand) that reflected the conversion of C’s to T’s in the LM libraries (**Table S1**). For example, in the plasma from patient INDI 995, a patient with stage IB ovarian clear cell adenocarcinoma, a *PIK3CA* p.H1047R mutation was found in 3.1% in the LA library. This mutation creates a CpG site (C<u>A</u>T->**C**<u>G</u>T) in the mutant molecules. To assess whether the mutant molecules found in LA molecules were methylated in the LM libraries, we amplified the LM libraries with primers in **Table S1**. Overall, 409 UIDs were found in both LM and LA libraries. Of those, 11 (2.7%) contained the *PIK3CA* p.H1047R mutation. The lower fraction of mutants detected (2.7% vs. 3.1%) is likely due to the distance of primers from the mutation of interest, such that some very short amplified DNA molecules (of the 409) did not span the mutation. As expected, a high fraction of mutant UIDs (82% of 11) were methylated in the **<u>C</u>G**T trimer, compared to only 1.3% of the 398 C residues in the analogous <u>C</u>AT trimer in the wildtype UIDs, presumably due to incomplete bisulfite conversion (**Fig. 7**).

**Fig. 7:**
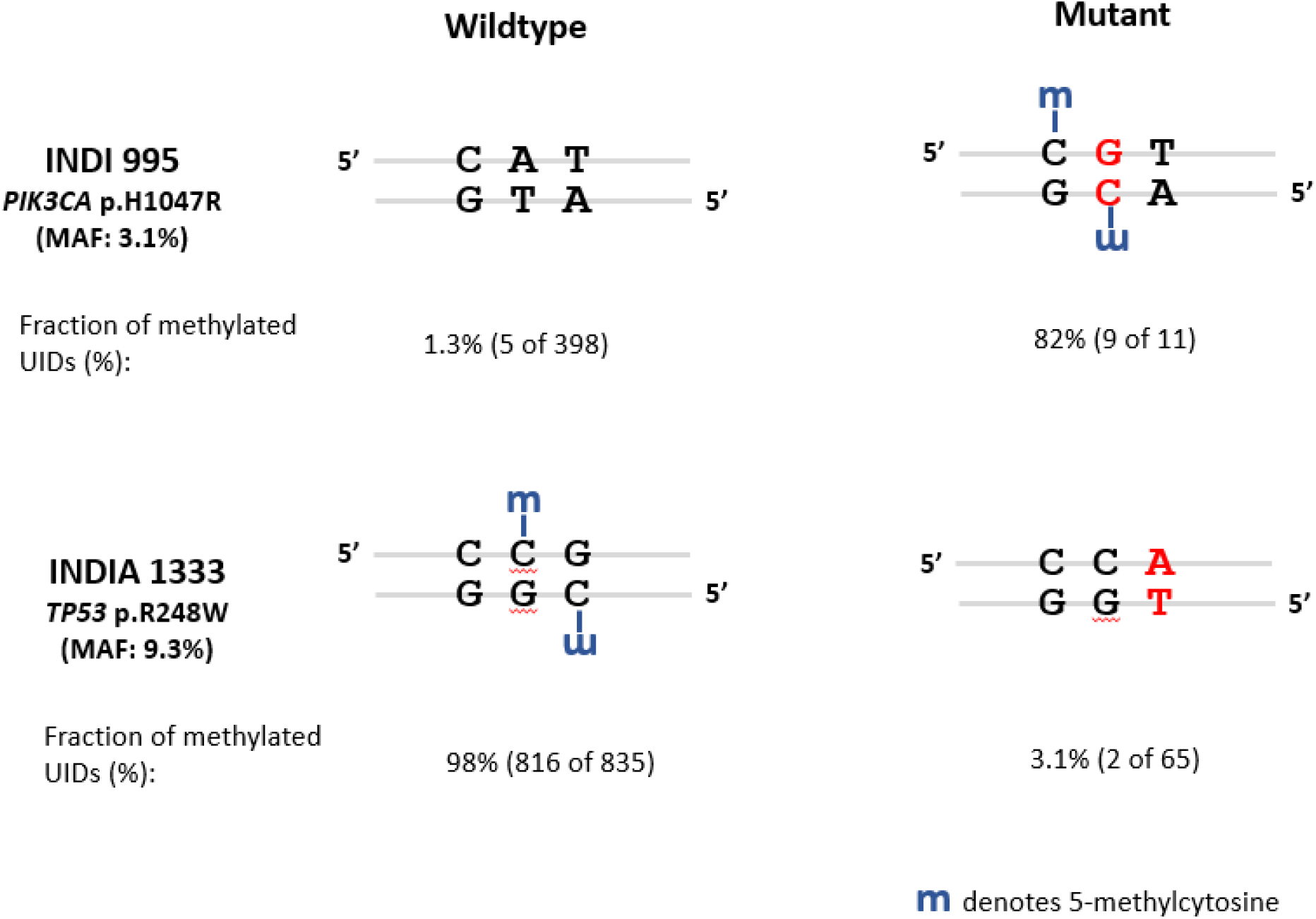
Determination of methylation status in wildtype and mutant DNA fragments in cfDNA. Two patient samples had mutations identified in the MethylSaferSeqS LA libraries that is expected to alter the methylation status of mutant molecules in the LM libraries. In INDI 955, the PIK3CA p.H1047R mutation was found at 3.1% of the UIDs in the MethylSaferSeqS LA library. This mutation creates a CpG site in mutant molecules. In INDIA 133, the TP53 p.R248W mutation was found in 9.3% of the UIDs in the MethylSaferSeqS LA library. This mutation erases a CpG site in mutant molecules. The expected methylation status of the wildtype and mutant molecules are shown for each patient, along with the fraction of methylated UIDs found for the wildtype and mutant molecules. MAF = mutant allele fraction.

Similarly, in another patient, INDIA 1333, who had stage IIB pancreatic adenocarcinoma, a *TP53* p.R248W mutation was found in the LA libraries at 9.3% MAF. This mutation occurred at a CpG site (C**C<u>G</u>**->CC<u>A</u>). As above, we performed targeted enrichment for this mutant position in the LM libraries with the primers listed in **Table S1**. There were 900 UIDs found in both LA and LM libraries. Of those, 65 UIDs (7.2%) contained the *TP53* p.R248W mutation. A high fraction of wildtype UIDs (98% of 835 UIDs) were methylated in the C**<u>C</u>G** trimer (**Fig. 7**). As expected, only 3.1% of the 65 C residues in the analogous C<u>C</u>A trimer in the mutant UIDs appeared to be methylated, presumably due to incomplete bisulfite conversion (**Fig. 7**).

## DISCUSSION

In the era of precision medicine, it would be useful to develop tools that make use of our burgeoning knowledge of the cancer genome and epigenome. Here we described a new method, MethylSaferSeqS, that can detect genetic and epigenetic changes in the same DNA molecules. Using plasma samples from cancer patients, we showed that the expected mutations, copy number changes, and methylation can be detected with MethylSaferSeqS. With simple modifications, MethylSaferSeqS can be adapted to any library preparation method compatible with next-generation sequencing, including those that support duplex sequencing, and can do so without greatly compromising the efficiency of molecule recovery.

There are a variety of potential applications for MethylSaferSeqS. One obvious application is to incorporate tumor-specific epigenetic and genetic markers for cancer detection. For this purpose, MethylSaferSeqS allows for concurrent construction of the libraries for epigenetic and genetic assessments, alleviating the need to split the sample for each assay and maximizing sensitivity. Furthermore, methylation markers can be tissue-specific as well as cancer-specific, such that they essentially provide fingerprints that can be used to trace the tissue of origin. Finally, MethylSaferSeqS libraries can obviously be used to study other features related to cancer, such as cell-free DNA fragmentomics, in the context of the genetic and epigenetic status of each DNA molecule.

There are several limitations to our study. First, we did not distinguish somatic mutations that were derived from neoplastic cells from those derived from hematopoietic cells. Thus we did not exclude mutations arising from clonal hematopoiesis of indeterminate potential (CHIP). In a diagnostic test, those mutations can be excluded by studying the DNA from matched white blood cells. This was not within the scope of our study, as our goal was to demonstrate that MethylSaferSeqS can reliably detect somatic mutations regardless of the source. However, given only one of the 67 healthy individuals (1.5%) had a detectable mutation, we do not believe that CHIP was a major source of mutations reported in our study.

Another limitation in our study was the loss of DNA during bisulfite treatment resulting in relatively low number of DNA molecules queried for methylation in LM libraries. Even with our modified protocol and reduced conversion time, a significant fraction of DNA (∼30%) was degraded during bisulfite treatment. To increase recovery further would require gentler treatment conditions that result in a lower conversion efficiency, which could lead to overestimation of the fraction of methylated molecules. In our study, a lower recovery did not affect our ability to perform shallow whole genome sequencing, given only a small fraction of molecules was sampled. To increase recovery, MethylSaferSeqS can be readily adapted to incorporate bisulfite-free methods for generating LM libraries (31). Depending on the analytical sensitivity required for the application, targeted enrichment approaches using capture or PCR may be necessary. The combination of higher recovery and enrichment would allow for duplex sequencing for methylation (i.e. presence of a methylated CpG dyad on both strands of the same molecule) for higher accuracy. However, duplex sequencing is not generally necessary for the evaluation of methylation in CpG-rich regions because errors from library construction or sequencing would likely affect only one rather than multiple CpGs.

Finally, we did not attempt to show that the concurrent evaluation of methylation, mutation, and copy number changes in the same DNA molecules could add sensitivity to existing detection methods. But with this first demonstration of a versatile method that is readily adaptable to existing sequencing and experimental strategies for the simultaneous detection of these features, our study sets the stage for a large prospective trial that can evaluate MethylSaferSeqS for clinical applications.

## MATERIALS AND METHODS

### Plasma Collection and DNA Collection

This study was approved by the Institutional Review Boards for Human Research at Johns Hopkins Medical Institutes in compliance with the Health Insurance Portability and Accountability Act. All the participants provided written informed consent in accordance with the principles of the Declaration of Helsinki. DNA was purified from ∼5 mL of plasma using a BioChain Cell-free DNA Extraction kit (Cat # K5011625). Some of the samples recorded in Table S7 have been used for purposes described elsewhere (22). All patients were de-identified and patient IDs are not known to anyone outside the research group.

### Library Preparation

Libraries were prepared as previously described(1) with the following modifications: Custom 3’ and 5’ adaptors containing 5-methylcytosines rather than unmethylated cytosines were used (**Table S1**) during ligation. The adaptor sequences otherwise are identical to what has been used for SaferSeqS. Following ligation, both strands of the templates were copied with a single, deoxyuridine-containing, dual-biotinylated primer at 5 uM targeting the 3’ adaptor (**Table S1**) for 3 PCR cycles with KAPA HiFi HotStart Uracil +ReadyMix Kit (Roche Diagnostics; cat # 7959052001). The resultant molecules (one original strand bound to a complementary copied strand from the last PCR cycle, and two additional copied strands from the first two PCR cycles) were bound to Streptavidin MyOne Dynabeads T1 (ThermoFisher; cat # 65601). Beads and bound DNA were heated to 75°C in 22.5 ul water for 2 minutes to separate the original strands (now in the supernatant) from the copied strands that remain attached to the beads. The beads were then incubated again in 22.5 uL of water at 75°C for 2 minutes, and the DNA in the supernatant combined with the DNA in the first supernatant (total of 45 uL).

For preparation of the LM library, the DNA from the supernatant was deaminated with bisulfite using reagents from the EZ DNA Methylation Kit (Zymo Research; cat # D5004) per manufacturer’s protocol with the following modifications: After denaturation, 100 uL of CT Conversion Reagent was added to each well and incubated in the dark for 3 hours at 50°C, followed by a 10 minute hold at 4 deg C. The conversion reaction was then added to the Silicon-A-Binding plate and mixed with 600 uL of M-Binding buffer. The plate was centrifuged at 2800 g for 5 min, and each column was subsequently washed with 500 uL of M-Wash buffer. After washing, 200 uL of M-Desulphonation buffer was added to each well and incubated at room temperature for 15 minutes. The plate was then centrifuged at 2800g for 5 minutes, and washed twice with 600 uL then 400 uL of M-Wash buffer. The DNA was finally eluted twice in 21 uL of M-elution buffer each time for a total volume of ∼42 uL, and amplified as below.

For preparation of the LA library, the copied strands bound to streptavidin beads were released by treatment with USER (Uracil-Specific Excision Reagent) enzyme (New England Biolabs; cat # M5505S). USER contains a mixture of Uracil DNA glycosylase and the DNA glycosylase-lyase Endonuclease VIII that in combination cleaves at the deoxyuridine base in the primer (**Fig. 1**). The Dynabeads with bound copied molecules were incubated with 3 uL of USER and 37 uL of Elution Buffer (Qiagen, Cat 19086) in a 40 uL reaction for 60 minutes at 37°C. Following incubation, the cleaved DNA strands were separated from the beads using a magnet, collected in the supernatant, and added to PCR reactions as described below.

For the amplification of LM libraries, ∼42 uL of purified DNA after bisulfite conversion was added to PCR reactions performed with KAPA HiFi HotStart Uracil +ReadyMix Kit (Roche Diagnostics; Cat # 7959052001) with primers at a final concentration of 5 uM (**Table S1**) in 100 uL reactions. For the amplification of LA libraries, 40 uL of DNA that was cleaved from the streptavidin beads was added to PCR reactions performed with KAPA HiFi HotStart +ReadyMix Kit (Roche Diagnostics; cat # K2601) with primers at a final concentration of 5 uM (**Table S1**) in 100 uL reactions. All libraries were amplified for 8 cycles as follows: 98°C for 45 seconds, then 8 cycles of 98°C for 15 seconds to denature, 60°C for 30 seconds to anneal, and 72°C for 30 seconds to extend. Amplified DNA from each reaction was purified using 180 uL SPRISelect beads (Beckman Coulter) and eluted using 100 uL of EB (Qiagen, cat # 19086). For WGS of either the LA or LM libraries, 100 ng of library DNA was amplified in 50 uL reactions in Ultra Q5 with primers at 2 uM (**Table S1**) for 7 cycles with the following conditions: 98C for 30 seconds, then 7 cycles of 98C for 10 seconds to denature and 65°C for 75 seconds to anneal and extend. For amplification of specific regions of the genome, anchored hemi-nested PCR for LM libraries were done using the primers described in **Table S1**. For amplification of the 48 regions in LA performed, primers were used as previously described (1).

### Sequencing and analysis

Barcoded libraries were sequenced using 100 bp paired-end runs (200 cycles) on a NovaSeq 6000 to an average depth of 75 million read pairs per sample. Mutation analysis was performed after demultiplexing, and grouping reads into duplex families, carried out as previously described (1). For methylation analysis, BSMAP was used to align each paired-end read to the bisulfite-converted hg19 genome, and the fractional methylation calculated with BSMAP’s methratio.py script (32). The intercept function in BEDtools (33) was then used to identify the fraction of methylation in the targeted regions (**Table S6**). The conversion efficiency for each sample was estimated from regions that are unmethylated such as in the 14 unmethylated bases in the UIDs (**Table S7**). For comparison of UIDs in the LA and LM libraries, all 14 bases in UIDs were presumed to undergo complete conversion during bisulfite treatment. The methylation fraction for each region was calculated from the total number of unconverted cytosines in CpG’s to the total number of CpGs per region:

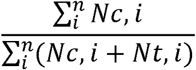

Where *n* represents the total number of CpG sites in the region, *i* represents a particular CpG site, *Nc,i* represents the number of unconverted C’s in the CpG site, and *Nt,i* represents the number of converted C to T’s in the CpG site.

This fraction was then adjusted to obtain values above the non-converted background and to account for minor differences in conversion efficiency across experiment as follows:

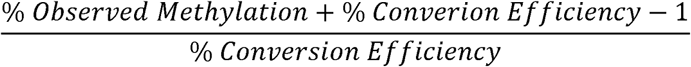

Only regions with more than 10 reads are included in **Table S7**. For cfDNA tissue deconvolution in plasma from healthy individuals using quadratic programming, the methods as previously described(21) were used with a total of 5,653 markers. For the identification of copy number changes, the reads from WGS of MethylSaferSeqS LA libraries and SaferSeqS libraries were processed using ichorCNA algorithm(34). As more stringent criteria, we required logR ratio > 0.16 (for copy number gain) or < -0.16 (for copy number loss) to filter out borderline regions, as well as number of bins per region > 20 (equivalent of 20 × 500,000 basepairs, or 10MB) to ensure that sufficiently large number of markers were used for mapping. Mutations are identified in the SaferSeqS and MethylSaferSeqS libraries using the following criteria: # Mutant UIDs (Supercalimutants) > 1, Family count within each UID family > 1, mutations that were either inactivating in tumor suppressors or present with > 5 counts in the Catalogue Of Somatic Mutations In Cancer Database (35). All sequencing data will be deposited in European Bioinformatics Institute EGAS00001006839.

## Data Availability

All sequencing data will be deposited in European Bioinformatics Institute EGAS00001006839.

## ACKNOWLEDGEMENT

We thank the individuals who participated in this study for their courage and generosity. We also thank Dr. S. Markowitz for insightful comments and review of this manuscript. We are grateful for C. Blair and K. Judge for expert technical and administrative assistance, and to E. Cook for illustrative assistance. This work was supported by The Lustgarten Foundation for Pancreatic Cancer Research, The Virginia and 297 D.K. Ludwig Fund for Cancer Research, The Conrad N. Hilton Foundation, The Sol Goldman Charitable Foundation, and National Institutes of Health grants (U01 CA200469, U01 CA62924, T32 GM136577, U01 CA06973, and T32 CA009071).

